# Lack of effectiveness of Bebtelovimab Monoclonal Antibody Among High-Risk Patients with SARS-Cov-2 Omicron During BA.2, BA.2.12.1 and BA.5 Subvariants Dominated Era

**DOI:** 10.1101/2022.12.06.22283183

**Authors:** Srilekha Sridhara, Ahmet B. Gungor, Halil K. Erol, Mohanad Al-Obaidi, Tirdad T. Zangeneh, Edward J. Bedrick, Venkatesh K. Ariyamuthu, Aneesha Shetty, Abd A. Qannus, Katherine Mendoza, Sangeetha Murugapandian, Gaurav Gupta, Bekir Tanriover

## Abstract

Severe acute respiratory syndrome coronavirus 2 (SARS-CoV-2) Omicron subvariants are expected to be resistant to Bebtelovimab (BEB) monoclonal antibody (MAb) and the real-world experience regarding its effectiveness is scarce. This retrospective cohort study reports a data analysis in Banner Healthcare System (a large not-for-profit organization) between 4/5/2022 and 8/1/2022 and included 19,778 Coronavirus disease-19 (COVID-19) positive (by PCR or direct antigen testing) patients who were selected from Cerner-Electronic Health Record after the exclusions criteria were met. The study index date for cohort was determined as the date of BEB MAb administration or the date of the first positive COVID-19 testing. The cohort consist of COVID-19 infected patients who received BEB MAb (N=1,091) compared to propensity score (PS) matched control (N=1,091). The primary outcome was the incidence of 30-day all-cause hospitalization and/or mortality. All statistical analyses were conducted on the paired (matched) dataset. For the primary outcome, the event counts and percentages were reported. Ninety-five percent Clopper-Pearson confidence intervals for percentages were computed. The study cohorts were 1:1 propensity matched without replacement across 26 covariates using an optimal matching algorithm that minimizes the sum of absolute pairwise distance across the matched sample after fitting and using logistic regression as the distance function. The pairs were matched exactly on patient vaccination status, BMI group, age group and diabetes status. Compared to the PS matched control group (2.6%; 95% confidence interval [CI]: 1.7%, 3.7%), BEB MAb use (2.2%; 95% CI: 1.4%, 3.3%) did not significantly reduce the incidence of the primary outcome (p=0.67). In the subgroup analysis, we observed similar no-difference trends regarding the primary outcomes for the propensity rematched BEB MAb treated and untreated groups, stratified by patient vaccination status, age (<65 years or ≥65), and immunocompromised status (patients with HIV/AIDS or solid organ transplants or malignancy including lymphoproliferative disorder). The number needed to treat (1/0.026-0.022) with BEB MAb was 250 to avoid one hospitalization and/or death over 30 days. The BEB MAb use lacked efficacy in patients with SARS-CoV-2 Omicron subvariants (mainly BA.2, BA.2.12.1, and BA.5) in the Banner Healthcare System in the Southwestern United States.

## INTRODUCTION

Severe acute respiratory syndrome coronavirus 2 (SARS-CoV-2) continues to evolve into new variants of concern (VOC) characterized by mainly spike receptor binding domain mutations, which are the target of authorized neutralizing monoclonal antibodies (MAb) to reduce hospitalization and death.(1) The spike protein mutations of SARS-CoV-2 Omicron subvariants have reduced susceptibility to earlier authorized MAbs (e.g. bamlanivimab-etesevimab, casirivimab-imdevimab, and sotravimab) for outpatient treatment of coronavirus disease-19 (COVID-19).(1-5) Based on invitro and limited clinical data(6), the Food and Drug Administration (FDA) granted Emergency Use Authorization (EUA) for LY-CoV1404 (Bebtelovimab [BEB]) on February 11, 2022, as an alternative therapy for high-risk patients with mild to moderate COVID-19.(4) BEB is an alternative treatment option for patients who are unable to receive remdesivir 3-days IV treatment due to logistic challenges or have contraindications for the use of nirmaltrevir/ritonavir due to severe drug-drug interactions. Bebtelovimab was recommended based on laboratory results indicating potent activity against the Omicron VOC and other VOCs based on data from the Phase 2 BLAZE-4 study.(4, 6) However, there is still no phase 3 clinical trial data to support BEB’s use and real-world experience is limited in the Omicron subvariants dominated era.(7, 8)

In this study, we assessed the composite outcome (all-cause hospitalization and/or death over 30-day) in high-risk outpatients, who received BEB MAb compared to the propensity score (PS) matched untreated control group for COVID-19 in the Banner Healthcare System (a large not-for-profit organization) in the Southwestern United States, during a period (4/5/2022-8/1/2022) dominated by SARS-CoV-2 Omicron BA.2, BA.2.12.1, and BA.5 subvariants.(9)

## METHODS

### Patient Consent Statement

This study was approved by the Institutional Review Board of the University of Arizona with a waiver of patient consent given the retrospective nature of the study. The study adhered to the Strengthening the Reporting of Observational Studies in Epidemiology (STROBE) statement (See Supplemental Document).

### Overview

This observational retrospective cohort study of positive COVID-19 patients was conducted between April 5, 2022, and August 1, 2022. Patients’ follow-up date was censored on August 31, 2022. All data pertaining to BEB MAb treated patients and untreated patients were captured from electronic health records (Cerner EHR) in the Banner Health Care System, which houses thirty hospitals and several clinics across the Southwestern United States, mainly in Arizona. A multidisciplinary team formed under the Banner Health Care System Monoclonal Antibody Treatment program reviews patients’ eligibility for antiviral therapy (remdesivir and nirmatrelvir-ritonavir as the first line agents) and an alternative MAb treatment, guided by the FDA EUA.(10) The alternative BEB MAb therapy (175 mg administered as a single intravenous injection over 30 seconds) is indicated for mild-to-moderate severe SARS-CoV-2 infection (within 7 days of symptom onset) in adults who are at high-risk for progression to severe disease and in children older than 12 years-old and weighing 40 kg or above.

In this study, 19,778 COVID-19 positive (by PCR or direct antigen testing) patients were selected from Cerner-EHR after exclusions were made (Figure 1). During the study period, there were 12 MAB infusion sites (for the treatment cohort) and 128 testing sites in the Banner Health Care System. The study index date for cohorts was determined as the date of BEB MAb administration or the date of the first positive COVID-19 testing. Patients were excluded if they were younger than 18 years of age, in hospice care, received BEB MAb in the inpatient setting, received tixagevimab-cilgavimab prophylactic MAb (Evusheld) within last 3 months/nirmatrelvir-ritonavir (Paxlovid) within 15 days/molnupiravir (Lagevrio) within 15 days of index date, or weighted less than 40 kilograms. The resulting pre-propensity matched study cohort comprised 1,099 BEB MAb treated patients and 18,679 untreated patients. Demographic and clinical covariates of both cohorts were extracted from the EHR. Clinical covariates were derived from the Charlson Comorbidity Index codes (based on International Classification of Diseases, Tenth Revision [ICD-10] codes documented in the EHR within five years preceding the patient index date). The post-propensity match cohort consisted of 1,091 pairs (N =2,182 patients).

**Figure 1.**
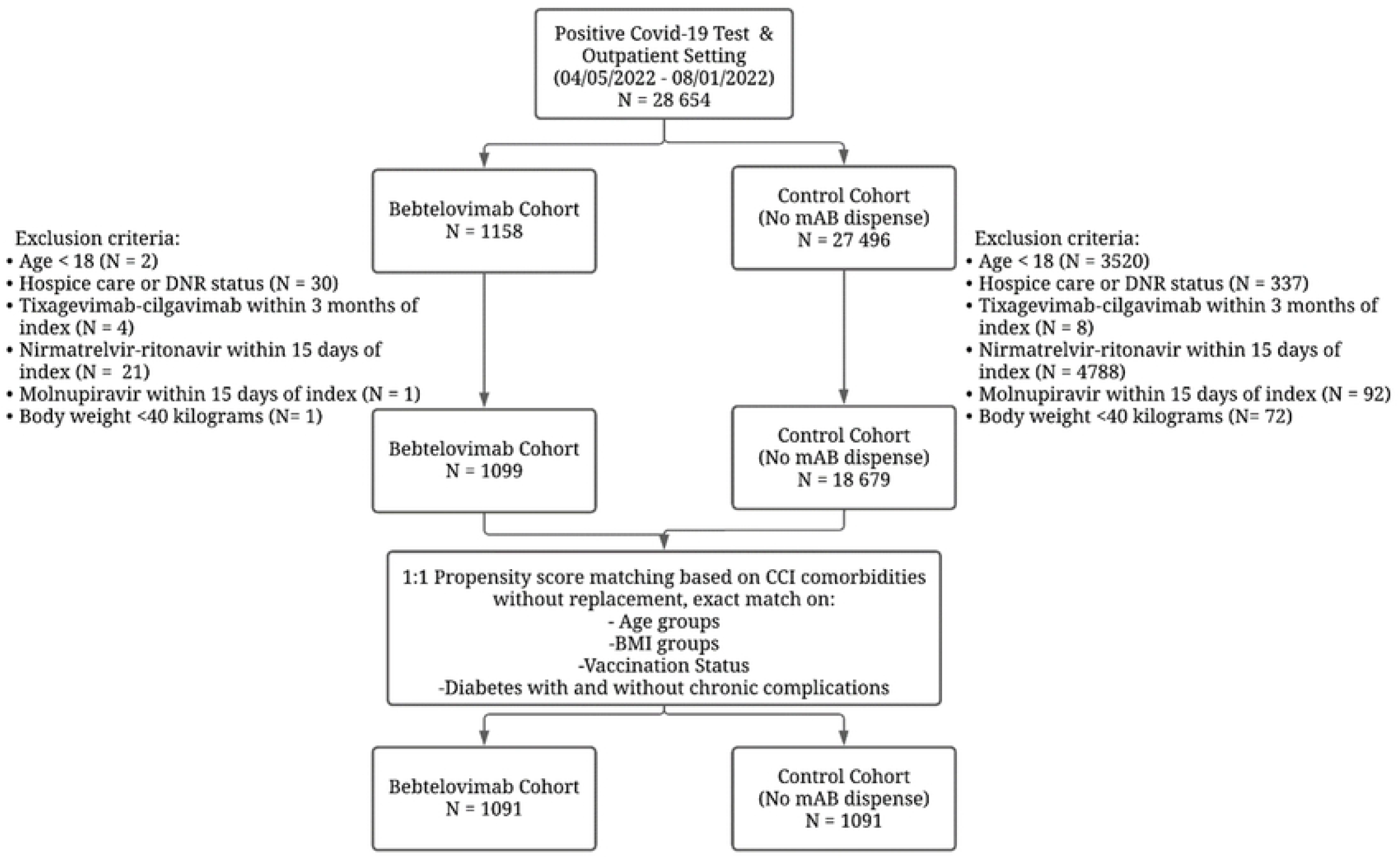
Study cohort selection.

### Outcome

The primary outcome was the incidence of all-cause hospitalization and/or mortality (the composite outcome) at 30-days of the index date in the post-propensity matched cohort.

### Statistical Methods

All statistical analyses were conducted on the paired (matched) dataset. For the primary outcome, the event count and percentage of the event was reported. Ninety-five percent Clopper-Pearson confidence intervals for percentages were computed in the R package Exactci. The study cohorts were 1:1 propensity matched without replacement across 26 covariates using an optimal matching algorithm that minimizes the sum of absolute pairwise distance across the matched sample after fitting and using logistic regression as the distance function. The pairs were matched exactly on patient vaccination status, BMI group, age group and diabetes status. Patients were classified as fully vaccinated if they had at least two or three (depending on immunocompromised status) COVID-19 mRNA technology vaccine (Pfizer or Moderna) reported in the EHR. The vaccination status of Arizona residents is available through a web-portal (the Arizona State Immunization Information System).(11)

The covariate balance was assessed by comparing pre- and post-match standardized mean differences (SMDs). MatchIt package from the statistical computing software R was used to build the propensity models. For each outcome, event count, percentage with the event and ninety-five percent confidence intervals have been reported. Exact McNemar’s test was used to compare the proportions in the pair dataset and the 95% confidence intervals for proportions were calculated. P-values <0.05 was considered statistically significant. The matched sets were constructed for the subgroup analysis and the incidence of the composite outcome was reported for the subgroups stratified by vaccination status (fully vaccinated and not fully vaccinated), age groups (age <65 and age ≥65), and immunocompromised status (patients with comorbidities including HIV/AIDS, malignancy and solid organ transplantation, and patients without these comorbidities). The Kaplan-Meier estimator was used to plot curves for the composite outcome between the post-PS matched groups during the study period. We fitted a multivariable Cox proportional hazard regression model predicting the composite outcome in the PS matched group.

## RESULTS

### Patient Characteristics

Table 1 shows the characteristics of BEB MAb and untreated control cohorts before and after propensity matching. All post-propensity matching covariate SMDs were < 0.1 threshold, indicating an optimal matching. In the post propensity matched cohort, the median age of patients in the BEB MAb treatment group was 64 (interquartile range [IQR], 50-74) years; 43% were male, and 78.7% were White race and 68.6% patients were fully vaccinated. Some of the high-risk characteristics included age ≥60 years (58.7%), hypertension [52.5%], diabetes mellitus (31.7%), chronic pulmonary disease (31.4%), BMI ≥35 kg/m2 (27.3%), chronic kidney disease–any stage (16.9%), chronic liver disease (13.8%), human immunodeficiency virus infection (HIV/AIDS) and/or opportunistic infections (11%), heart failure (8.3%), malignancy including lymphoproliferative disease (7.7%), and solid organ transplant and hematopoietic stem cell transplants (4.9%).

**Table 1:**
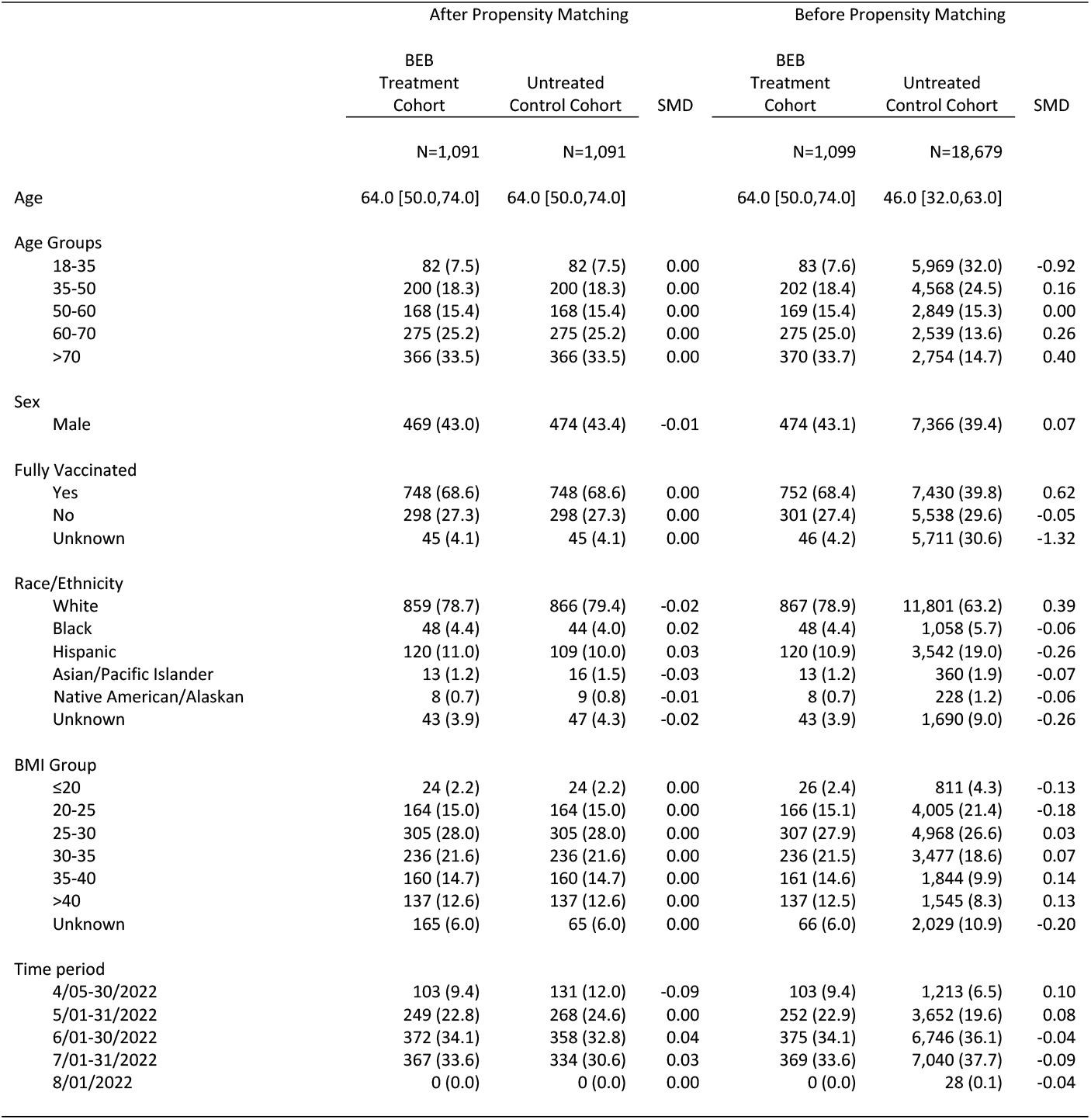
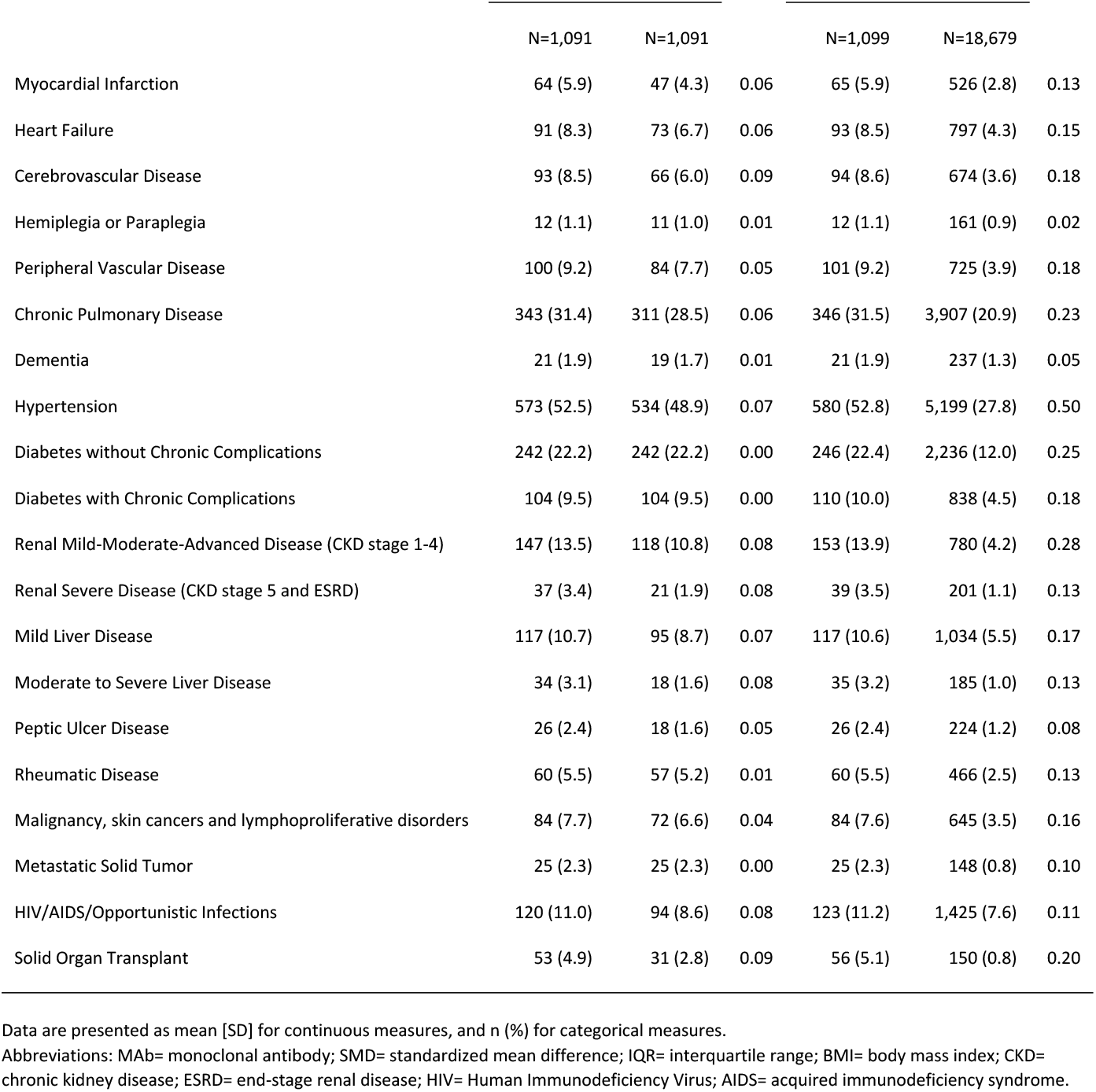
Patient characteristics and covariate balance before and after propensity matching.

### Outcomes

The incidence of the composite outcome in the pre-PS matched untreated control cohort was 1.8% (data not shown). Table 2 shows the result of the composite outcome within 30 days in the post propensity matched cohorts. Compared to the untreated control group, the incidence of patients with the composite outcome in the BEB MAb treated group within 30 days is 2.2% (95%: CI 1.4% to 3.3%) vs. 2.6% (95% CI: 1.7% to 3.7%) (P-value =0.67). The all-cause hospitalizations within 30 days in the BEB MAb cohort was 2.2% (95% CI: 1.4% to 3.3%) vs 2.5% (95% CI, 1.6% to 3.6%) (P value =0.77); the proportion of patients with all-cause mortality within 30 days was 0% (95% CI, 0% to 0%) vs 0.3% (95% CI, 0.1% to 0.8%; P-value =0.25). Figure 2 showed no difference between the Kaplan Meier curves for the composite outcome stratified by BEB MAb treatment status at last follow-up (P-value =0.27). The number needed to treat (1/0.026-0.022) with BEB MAb was 250 to avoid one hospitalization and/or death over 30 days.

**Table 2:** The primary composite outcome between the propensity matched Bebtelovimab (BEB) monoclonal antibody (MAb) and untreated control groups.

| Primary outcomes in post-propensity score-matched cohorts |  |  |  |  |  |  |
| --- | --- | --- | --- | --- | --- | --- |
|  | BEB MAb Treatment Group |  | Untreated Control Group |  | Difference in % with 95% CI** | P-value |
|  | N (%) | 95% CI* | N (%) | 95% CI* |  |  |
| Composite outcome within 30 days |  |  |  |  |  |  |
| Whole cohort | 24 (2.2) | 1.4, 3.3 | 28 (2.6) | 1.7, 3.7 | -0.4 (-1.7, 1.0) | 0.67 |
| All-cause hospitalization within 30 days |  |  |  |  |  |  |
| Whole cohort | 24 (2.2) | 1.4, 3.3 | 27 (2.5) | 1.6, 3.6 | -0.3 (-1.6, 1.1) | 0.77 |
| Mortality within 30 days |  |  |  |  |  |  |
| Whole cohort | 0 (0.0) | 0.0, 0.3 | 3 (0.3) | 0.1, 0.8 | -0.3 (-0.8, 0.1) | 0.25 |

Abbreviations: BEB= Bebtelovimab; MAb= monoclonal antibody; CI= Confidence Interval.
\* The Clopper-Pearson method was used to calculate 95% confidence intervals for the outcome percentages using the R package (Exactci).
\*\* CI for difference in paired proportions between the treatment and control cohorts.

**Figure 2.**
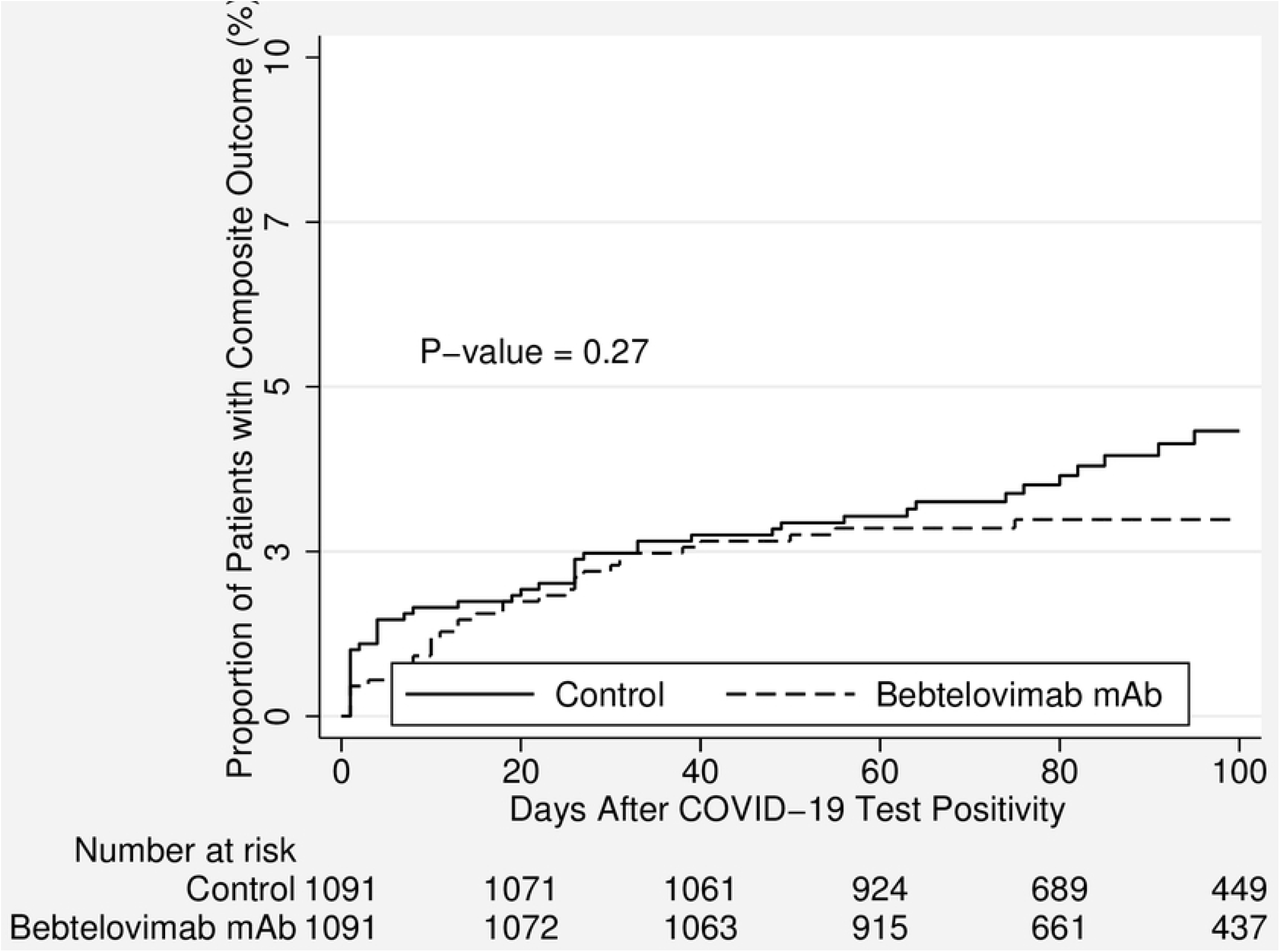
Kaplan Meier curves for the composite outcome in patients who received Bebtelovimab monoclonal antibody treated group vs. not-treated control group between April 5, 2022, and August 1, 2022.

In the subgroup analysis, we observed similar no-difference trends regarding the primary outcomes for the propensity rematched BEB MAb treated and untreated groups, stratified by patient vaccination status, age (<65 years or ≥65), and immunocompromised status (patients with HIV/AIDS or solid organ transplants or malignancy including lymphoproliferative disorder), see Table 3 below.

**Table 3:** Subgroup analysis for the primary composite outcome stratified by patient vaccination status (fully vaccinated vs. not fully vaccinated), age category (age <65 vs. age ≥65), and immunocompromised status (comorbidities including HIV/AIDS or solid organ transplants or malignancy) between the propensity matched Bebtelovimab (BEB) and untreated control groups.

| Primary outcome in the post-propensity-matched study cohort with subgroups |  |  |  |  |  |  |
| --- | --- | --- | --- | --- | --- | --- |
|  | BEB MAb Treatment Group |  | Untreated Control Group |  | Difference in % with 95% CI*** | P-value |
|  | N (%) | 95% CI* | N (%) | 95% CI** |  |  |
| Fully vaccinated*<br>N=1,496 | 7 (0.9) | 0.4, 1.9 | 11 (1.5) | 0.7, 2.6 | -0.5 (-1.8, 0.7) | 0.48 |
| Not fully vaccinated<br>N=596 | 17 (5.7) | 3.4, 9.0 | 17 (5.7) | 3.4, 9.0 | 0.0 (-4.0, 4.0) | 1.00 |
| Immunocompromised<br>N=250 | 9 (7.2) | 3.3, 13.2 | 11 (8.8) | 4.5, 15.2 | -1.6 (-9.2, 6.0) | 0.81 |
| Not-Immunocompromised<br>N=1,636 | 10 (1.2) | 0.6, 2.2 | 7 (0.9) | 0.3, 1.8 | 0.4 (-0.8, 1.5) | 0.63 |
| Age ≥65<br>N=1,014 | 13 (2.6) | 1.4, 4.3 | 17 (3.4) | 2.0, 5.3 | -0.8 (-3.1, 1.5) | 0.57 |
| Age <65<br>N=1,068 | 8 (1.5) | 0.6, 2.9 | 9 (1.7) | 0.8, 3.2 | -0.2 (-2.0, 1.6) | 1.00 |

Abbreviations: HIV= human immunodeficiency virus; AIDS= acquired immunodeficiency syndrome; Bebtelovimab= BEB; MAb= monoclonal antibody; CI= Confidence Interval.
\*This analysis included the patients with known vaccination status only.
\*\*The Clopper-Pearson method was used to calculate 95% confidence intervals (CI) for the outcome percentages using the R package (Exactci).
\*\*\* CI for difference in paired proportions between the BEB MAb treatment and control cohorts.

### Hazard model for Composite Outcome among the Propensity Matched SARS-Cov-2 Infected Patients

Table 4 shows the multivariable Cox proportional hazards model, (accounted for the paired data) predicting hazards for the composite outcome among the post-PS patients. The BEB MAb use was not associated with statistically significant lower hazards of composite outcome (hazard ratio [HR] 0.75; 95% CI: 0.43 to 1.31, P-value =0.31). However, fully vaccinated status continued to be protective while age >65 and immunosuppressed status increased the hazards for primary outcome two to four folds, respectively.

**Table 4.** Multivariable Cox proportional hazard model* for the composite outcome among the post-propensity matched COVID-19 infected patients in the Banner Healthcare System between April 5, 2022, and August 1, 2022.

|  | Hazard Ratio (95%<br>Confidence Interval) | Standard<br>Error | P-value |
| --- | --- | --- | --- |
| Bebtelovimab monoclonal<br>antibody use (yes vs. no) | 0.75 (0.43-1.31) | 0.21 | 0.31 |
| Fully vaccinated status<br>(Yes vs. no) | 0.23 (0.12-.42) | 0.07 | <0.001 |
| Age (≥65 vs. <65) | 2.07 (1.15-3.74) | 2.41 | 0.02 |
| Immunocompromised**<br>(Yes vs. no) | 4.60 (2.58-8.19) | 5.19 | <0.001 |

\*Accounted for the paired data.
\*\*Immunocompromised status (the patients with HIV/AIDS or solid organ transplants or malignancy including lymphoproliferative disorder).

## DISCUSSION

In this retrospective propensity matched analysis, the incidence of the composite outcome was low (2.2%-2.6%) and treatment with the BEB MAb lacked efficacy against SARS-CoV-2 Omicron during an era dominated by BA.2, BA.2.12.1, and BA.5 subvariants to reduce the all-cause hospitalization and mortality over 30 days in Banner Health Care System in the Southwestern United States. Moreover, in the subgroup analysis for the composite outcome stratified by patient vaccination status, age category, and immunocompromised status between the PS matched groups, BEB MAb use failed to show significant efficacy. The hazards for the composite outcome were lower in the BEB MAb group but not statistically significant. However, fully vaccinated status continued to be protective while age >65 and immunosuppressed status increased the hazards for primary outcome two to four folds, respectively. Similar finding from epidemiological study showing possible protective immunity from previous infections and vaccinations, and that older age can result in worse outcomes during the Omicron wave.(12) Such findings can help stratifying risk groups when administering COVID-19 therapeutics.

The only published (non-peer-reviewed data) on the efficacy of BEB MAb comes from the Phase II Blaze 4 clinical trial during the period of alpha and delta waves, which showed that the incidence of the primary outcome (hospitalization or death over 29 days) in the BEB MAb arm compared with the control arm was similar, around 3%.(6) While in vitro studies showing preserved neutralization of SARS-COV2 variants (BA.2 and BA. 5), (13, 14) it was not demonstrated in clinical trials. Hence, the real-world experience with BEB Mab use, especially during the periods of new variants emergence, is limited to a couple of recently published studies in the general population(7, 8) and solid organ transplant cohorts.(15, 16) A Mayo clinic study (N=2,833) reported that the BEB MAb use was associated with very low incidence of the primary outcome (1.4%, 95% CI: 1.2% to 1.7%) between 3/20/2022 and 6/14/2022, dominated by Omicron BA.2 subvariant.(8) However, the study was limited by a population of predominantly White and fully vaccinated (>90%) patients, and moreover, the study lacked a matched control group and did not clearly define exclusion criteria (e.g., tixagevimab-cilgavimab prophylactic MAb (Evusheld) use etc.). Therefore, in the absence of control group comparison, it is difficult to ascertain the author’s conclusion of primary outcome of 1.4% because of the fully vaccinated status of patients or the effect of the BEB MAb use. In contrast, our data suggest that the fully vaccinated patients had similar primary outcome of 0.9% in BEB MAb group vs. 1.5% in non-treated PS matched group, which signifies the importance of population immunity. In another study (N = 930 patients in each arm), the University of Pittsburgh researchers showed that BEB MAb use, between 3/30/2022 and 5/30/2022, significantly reduced 30-day hospitalization and/or death compared to the PS matched cohort, 3.1% vs. 5.5%, respectively.(7) But the protective effect was the most prominent among older, immunosuppressed and fully vaccinated patients. The authors did not exclude the patients who received tixagevimab-cilgavimab prophylactic MAb (Evusheld) and the cohort included small proportions of racial minorities (Blacks/Hispanics/Asians) comprised approximately 4% of the final study cohort. The SARS-Cov-2 Omicron BA.2 subvariant dominated the COVID-19 infections during that study period. In terms of SOT recipients who received the BEB MAb, another study from the Mayo Clinic(16) reported 3.3% incidence of the primary outcome in their cohort (N=92) and a smaller number of SOT recipients received BEB MAb compared to Sotrovimab MAb group. In contrast, the SOT recipients in our cohort (n=53) had much higher incidence of hospitalization and death (9.4% in BEB MAb group vs. 6.5% in control arm, P-value =0.64), data not shown. The differences in vaccination rates and the different effects of Omicron subvariants may account for this variation.

On November 10, 2022, the NIH COVID-19 Treatment Guidelines Panel(17) reported that certain rapidly increasing Omicron subvariants (e.g., BQ.1 and BQ.1.1) are likely to be resistant to BEB MAb(18) based on in vitro neutralization studies.(19) The panel recommend BEB MAb only as an alternative treatment for when preferred ritonavir-boosted nirmatrelvir or remdesivir are not available or contraindicated, and when the majority of spreading (>50%) Omicron subvariants in a given region are susceptible, the rating of evidence rated as level III (expert opinion). Later, on November 30, 2022, the U.S. Food & Drug Administration removed the EUA of BEB due to the fact that a non-susceptible SARS-CoV-2 subvariants account for majority of COVID 19 cases (Omicron BQ.1 and BQ.1.1 subvariants infections to be above 57% nationally).(20)

Large observational (real-life experiences) studies are necessary to show the efficacy assessment of MAbs in the setting of continuously mutating SARS-Cov-2 when conducting conventional randomized controlled clinical trials may not be practical. However, our study has several limitations: 1) retrospective study design not allowing to rule residual confounding; 2) lack of symptom severity assessment among patients (possibility of more symptomatic patients on the BEB MAb group vs. asymptomatic patients on the control arm); 3) not measuring the impact of immunity through prior COVID-19 infection(s); 4) not knowing patients’ vaccination booster status (3^rd^ or 4^th^ booster); 5) lack of specific SARS-Cov-2 Omicron subvariant genotype sampling; 6) not capturing patients may have received Ritonavir-boosted nirmatrelvir (Paxlovid) and other approved therapies by healthcare providers outside our healthcare system.

In conclusion, the BEB MAb use lacked efficacy in patients with SARS-CoV-2 Omicron subvariants in the Banner Healthcare System (a large not-for-profit organization) in the Southwestern United States. Under the light of the current study findings and an expectation of the majority of Omicron subvariants becoming resistant, the continuing use of BEB MAb may no longer be justified. Continuing real-world research from other large healthcare organizations in the different regions of the United States would be needed to assess generalizability.

## Data Availability

Data cannot be shared publicly because of HIPAA rules. Data are available from the The University of Arizona/Banner University Medical Group Institutional Data Access / Ethics Committee (contact Dr Bekir Tanriover via) for researchers who meet the criteria for access to confidential data.

## Acknowledgment

The authors thank to Chad Whelan, MD, Joshua Lee, MD, and Gordon Carr, MD, for their support.

## Conflict of Interest

The authors declared no conflict of interest related to this research. Dr. M Al-Obaidi reported that he received an honorarium from Shionogi Inc. and La Jolla pharmaceuticals for serving in their advisory board meetings.

## Funding/Support

None.

## Notes

### Competing Interest Statement

The authors have declared no competing interest.

### Author Declarations

This study was approved by the Institutional Review Board of the University of Arizona with a waiver of patient consent given the retrospective nature of the study. The study adhered to the Strengthening the Reporting of Observational Studies in Epidemiology (STROBE) statement.

## References

1. Iketani S, Liu L, Guo Y, Liu L, Chan JF, Huang Y, et al. Antibody evasion properties of SARS-CoV-2 Omicron sublineages. Nature. 2022;604(7906):553–6.

2. Takashita E, Yamayoshi S, Simon V, van Bakel H, Sordillo EM, Pekosz A, et al. Efficacy of Antibodies and Antiviral Drugs against Omicron BA.2.12.1, BA.4, and BA.5 Subvariants. N Engl J Med. 2022;387(5):468–70.

3. Bruel T, Hadjadj J, Maes P, Planas D, Seve A, Staropoli I, et al. Serum neutralization of SARS-CoV-2 Omicron sublineages BA.1 and BA.2 in patients receiving monoclonal antibodies. Nat Med. 2022;28(6):1297–302.

4. US Food and Drug Administration. COVID-19 update: FDA authorizes new monoclonal antibody for treatment of COVID-19 that retains activity against Omicron variant. Available at: https://www.fda.gov/news-events/press-announcements/coronavirus-covid-19-update-fdaauthorizes-new-monoclonal-antibody-treatment-covid-19-retains. Accessed on November 11, 2022.

5. Gershengorn HB, Patel S, Ferreira T, Das S, Parekh DJ, Shukla B. The clinical effectiveness of REGEN-COV in SARS-CoV-2 infection with Omicron versus Delta variants. PLoS One. 2022;17(12):e0278770.

6. Dougan M, Azizad M, Chen P, et al. Bebtelovimab, alone or together with bamlanivimab and etesevimab, as a broadly neutralizing monoclonal antibody treatment for mild to moderate, ambulatory COVID-19. medRxiv. Preprint and has not been peer-reviewed. Available at doi: https://doi.org/10.1101/2022.03.10.22272100.

7. McCreary EK, Kip KE, Collins K, Minnier TE, Snyder GM, Steiner A, et al. Evaluation of Bebtelovimab for Treatment of Covid-19 During the SARS-CoV-2 Omicron Variant Era. Open Forum Infect Dis. 2022;9(10):ofac517.

8. Razonable RR, O’Horo JC, Hanson SN, Arndt RF, Speicher LL, Seville TA, et al. Outcomes of Bebtelovimab Treatment is Comparable to Ritonavir-boosted Nirmatrelvir among High-Risk Patients with Coronavirus Disease-2019 during SARS-CoV-2 BA.2 Omicron Epoch. J Infect Dis. 2022.

9. CoVariants. Available at https://covariants.org/, accessed on 11/10/2022.

10. The Banner Health COVID-19 Treatment. Available at https://www.bannerhealth.com/staying-well/health-and-wellness/wellness/covid/treatment, accessed on 11/10/2022.

11. Arizona State Immunization Information System [ASIIS], available at https://asiis.azdhs.gov/, accessed on 11/12/2022.

12. Adjei S, Hong K, Molinari NM, Bull-Otterson L, Ajani UA, Gundlapalli AV, et al. Mortality Risk Among Patients Hospitalized Primarily for COVID-19 During the Omicron and Delta Variant Pandemic Periods - United States, April 2020-June 2022. MMWR Morb Mortal Wkly Rep. 2022;71(37):1182–9.

13. Cao Y, Yisimayi A, Jian F, Song W, Xiao T, Wang L, et al. BA.2.12.1, BA.4 and BA.5 escape antibodies elicited by Omicron infection. Nature. 2022;608(7923):593–602.

14. Westendorf K, Zentelis S, Wang L, Foster D, Vaillancourt P, Wiggin M, et al. LY-CoV1404 (bebtelovimab) potently neutralizes SARS-CoV-2 variants. Cell Rep. 2022;39(7):110812.

15. Shertel T, Lange NW, Salerno DM, Hedvat J, Jennings DL, Choe JY, et al. Bebtelovimab for Treatment of COVID-19 in Ambulatory Solid Organ Transplant Recipients. Transplantation. 2022;106(10):e463–e4.

16. Yetmar ZA, Beam E, O’Horo JC, Seville MT, Brumble L, Ganesh R, et al. Outcomes of bebtelovimab and sotrovimab treatment of solid organ transplant recipients with mild-to-moderate coronavirus disease 2019 during the Omicron epoch. Transpl Infect Dis. 2022;24(4):e13901.

17. The NIH COVID-19 Treatment Guidelines Panel’s Statement on Omicron Subvariants, Pre-Exposure Prophylaxis, and Therapeutic Management of Nonhospitalized Patients With COVID-19. Available at: https://www.covid19treatmentguidelines.nih.gov/therapies/statement-on-omicron-subvariants/. Accessed on November 20, 2022.

18. Food and Drug Administration. Fact sheet for healthcare providers: Emergency Use Authorization for bebtelovimab. 2022. Available at: https://www.fda.gov/media/156152/download. Accessed on November 20, 2022.

19. Cao Y, Jian F, Wang J, et al. Imprinted SARS-CoV-2 humoral immunity induces convergent Omicron RBD evolution. bioRxiv. 2022;Preprint. Available at: https://www.biorxiv.org/content/10.1101/2022.09.15.507787v4. Accessed on November 20, 2022.

20. FDA Announces Bebtelovimab is Not Currently Authorized in Any US Region. Avaiable at: https://www.fda.gov/drugs/drug-safety-and-availability/fda-announces-bebtelovimab-not-currently-authorized-any-us-region, accessed on December 3, 2022.

